# Identification of novel mechanistically distinct endotypes of pandemic influenza-associated acute respiratory failure

**DOI:** 10.1101/2020.07.12.20150474

**Authors:** Romit Samanta, Jake Dunning, Adam Taylor, Andrew I. Bayliffe, Rachel C. Chambers, Edwin R. Chilvers, Peter JM. Openshaw, Charlotte Summers, on behalf of the MOSAIC and EMINENT-ARDS investigators.

## Abstract

Respiratory viral pandemics result in large numbers of cases of acute respiratory failure arising from a single etiology, thus reducing the heterogeneity of precepting insult and allowing improved insight into the variation of host responses. In 2009-2011, an influenza pandemic occurred, with pH1N1 infecting millions of people worldwide. Here, we have used novel bioinformatic methods to combine clinical, protein biomarker, and genomic data from patients with influenza-associated acute respiratory failure to identify three mechanistically discrete sub-types with significantly different clinical outcomes. The three endotypes identified can be described as “neutrophil-driven” (16.3%), “adaptive” (51.9%), and “endothelial leak” (31.7%). The neutrophil driven patients display evidence of innate immune activation with associated multi-organ dysfunction and reduced 30-day survival. These patients could be differentiated from the adaptive endotype by an alteration in the GAIT-mechanism, a late transcriptional regulatory response to IFN-γ that acts to suppress innate immunity by reducing caeruloplasmin mRNA translation. Patients with the neutrophil-driven endotype had significantly increased IFN-γ levels but appeared unable to suppress their innate immune response. The endothelial leak endotype could be distinguished from both the neutrophil driven and adaptive endotypes by alterations in Slit-Robo signalling, a pathway important in the maintenance of endothelial barrier integrity; Although patients with this endotype required mechanical ventilation, they did not develop multi-organ failure in the manner of the neutrophil-driven endotype patients, and had significantly better clinical outcomes. Importantly, the endotypes identified were stable over 48 hours opening up the possibility of stratified interventional clinical trials in the future.

**One Sentence Summary:** We have identified three new mechanistically distinct subtypes of influenza associated acute respiratory failure, with differential clinical outcomes.

## Introduction

Acute respiratory distress syndrome (ARDS) is a heterogenous clinical syndrome characterized by acute refractory hypoxemia and bilateral pulmonary infiltrates on thoracic radiography. Patients with ARDS have a hospital mortality of 40-48% *(1, 2)*, but the wide variation in both the precipitating etiology and host responses have made understanding the mechanisms underlying ARDS challenging. This heterogeneity may explain why there is no licensed pharmacotherapy for this devastating but common condition.

Interest has developed across a number of clinical fields, to determine whether distinct sub-groups or endotypes, exist within heterogenous clinical syndromes such as asthma, chronic obstructive pulmonary disease, cancer, and more latterly ARDS*(3–6)*. Detailed biological data can now be combined with symptomatology and treatment information into models that identify patient clusters that might not otherwise have been apparent. The characteristics that these subgroups share are enriched for mechanisms of disease, rather than grouping patients based on observable features alone (disease phenotypes). For example, Hinks et al have shown that whilst clinically based phenotypes fail to stratify patients with asthma, mechanistically plausible endotypes can be identified *(3)*.

Calfee and colleagues demonstrated using retrospective latent class analysis (LCA) of data obtained during randomized controlled trials, that two endotypes of ARDS can be defined, described as hyperinflammatory and hypo-inflammatory *(7)*. The hyperinflammatory endotype (one third of the patients) was characterized by increased serum IL-6, soluble TNF receptor-1, and plasminogen activator inhibitor-1 concentrations, and decreased serum bicarbonate and platelet concentrations. Importantly, the degree of respiratory failure (used clinically categorize ARDS), did not differ between the endotypes. The identified latent classes have also been demonstrated in retrospective analyses of some but not all other clinical studies *(8)*, and the two classes have been shown to have differential treatment responses to interventions such as positive end expiratory pressure, intravenous fluids, and statin therapy *(7, 9, 10)*. Although LCA provides some mechanistic insights, it is not a complete model. These *post hoc* analyses have also only been able to include data from a limited number of protein biomarkers and clinical features assessed as part of the randomized controlled trials, limiting the insights they are able to gain from a heterogenous population of patients with ARDS.

Respiratory viral pandemics result in large number of cases of acute respiratory failure arising from a single etiology, thus reducing the heterogeneity of precepting insult and allowing improved insight into the variation of host responses. In 2009-2011, an influenza pandemic occurred, with pH1N1 infecting millions of people worldwide. In response to this, the Mechanisms of Severe Acute Influenza Consortium (MOSAIC) was established to characterize patients hospitalized in England with suspected pH1N1. Further detail on the study undertaken can be found in *(11)*.

Here, for the first time, we have used novel bioinformatic methods to combine clinical, protein biomarker, and genomic data from patients with influenza-associated acute respiratory failure to identify three mechanistically discrete sub-types, with significantly different clinical outcomes.

## Results

### Patient Characteristics

The MOSAIC study recruited 212 patients with confirmed influenza infection, of which 157 adults had serum biomarkers measured and 179 had whole blood gene expression determined by microarray. We restricted our analyses to patients with moderate/severe influenza confirmed on polymerase chain reaction (PCR), who had a respiratory sequential organ failure (rSOFA) score greater than or equal to two. Patients with rSOFA of two are defined as receiving oxygen therapy, whilst those with a score of three or four are defined as requiring mechanical ventilation. 104 patients with moderate/severe influenza had both serum biomarker and gene expression data available. There were no differences between baseline patient characteristics between those receiving oxygen therapy alone and those requiring mechanical ventilation (Table 1).

**Table 1:**
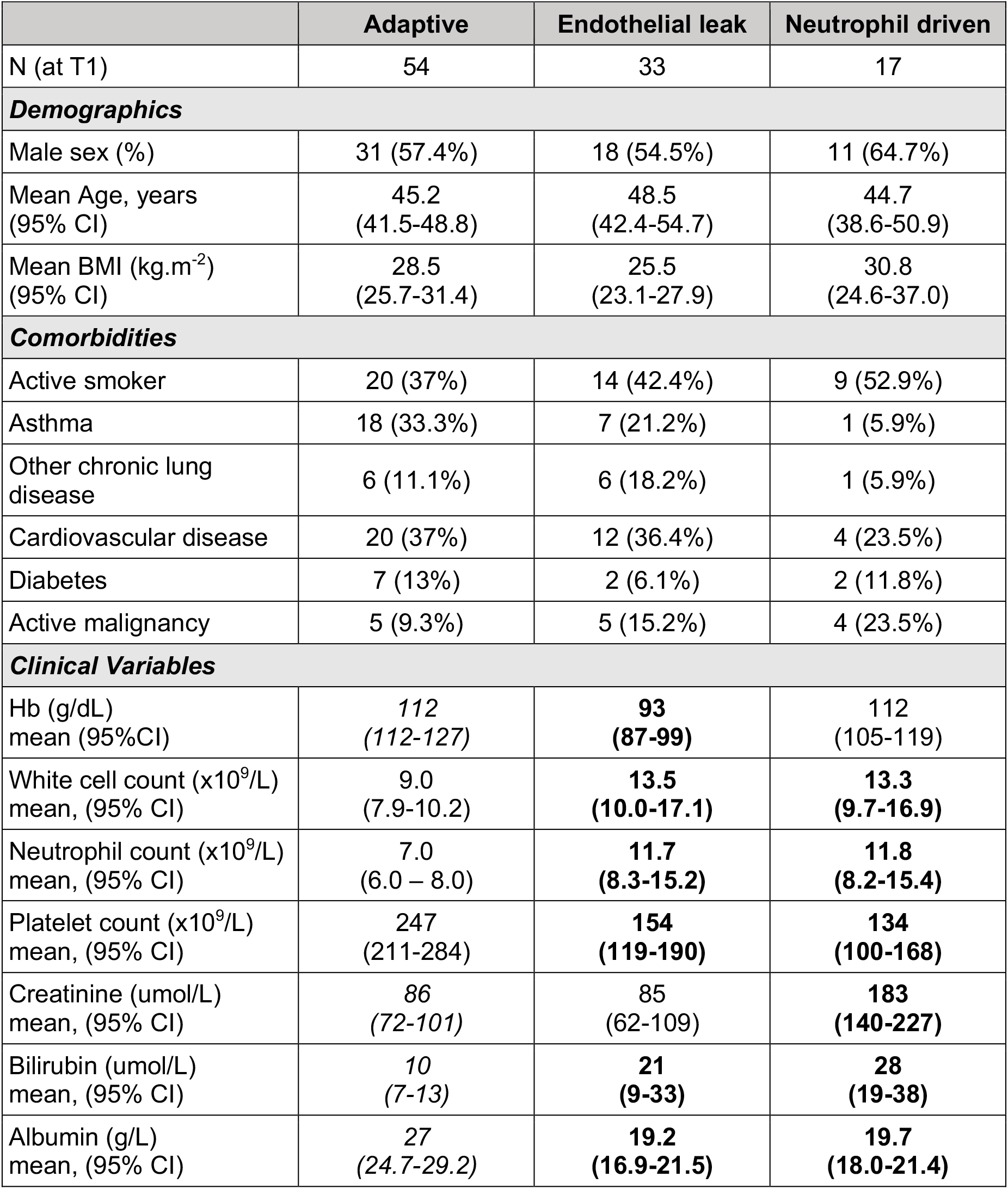
Endotype characteristics by demographics, past medical history and measured clinical variables. Categorical variables were tested by logistic regression. Continuous variables were compared using multiple-comparison corrected t-tests. For significantly different variables between endotypes, the reference value is in italics and the statistically significant value highlighted in bold face.

### Hierarchical clustering identifies three clusters of cytokine response in patients with moderate/severe influenza

The serum biomarker concentrations were log transformed and normalized prior to agglomerative, hierarchical, Ward clustering. The optimum cluster number was calculated by using both a k-means based elbow method and consensus methods using the *NbClust* R package (Figure S1) *(12)*. The optimum cluster number using these methods was calculated to be three, and there were 33 (cluster 1), 54 (cluster 2) and 17 (cluster 3) patients in each cluster, respectively.

Average serum biomarker concentrations in each patient cluster and heatmaps showed cluster 2 to have lower concentrations of cytokines associated with innate immune activation (such as TNFα and IL-6). Cluster 3 was characterized by high TNFα, IL-6, IL-15 and procalcitonin, whilst cluster 1 had moderately raised levels of these three mediators as well as increased concentrations of TNFR1 and TNFR2 (Figure 2 panel A). Patients in clusters 1 (48.5%) and 3 (64.7%) were significantly more likely to require mechanical ventilation compared with patients in cluster 2 (24.1%), suggesting that there were two sub-groups of patients requiring mechanical ventilation, with different biomarker profiles. There was no significant difference in viral load between patients in the three clusters (Figure S2).

**Figure 1:**
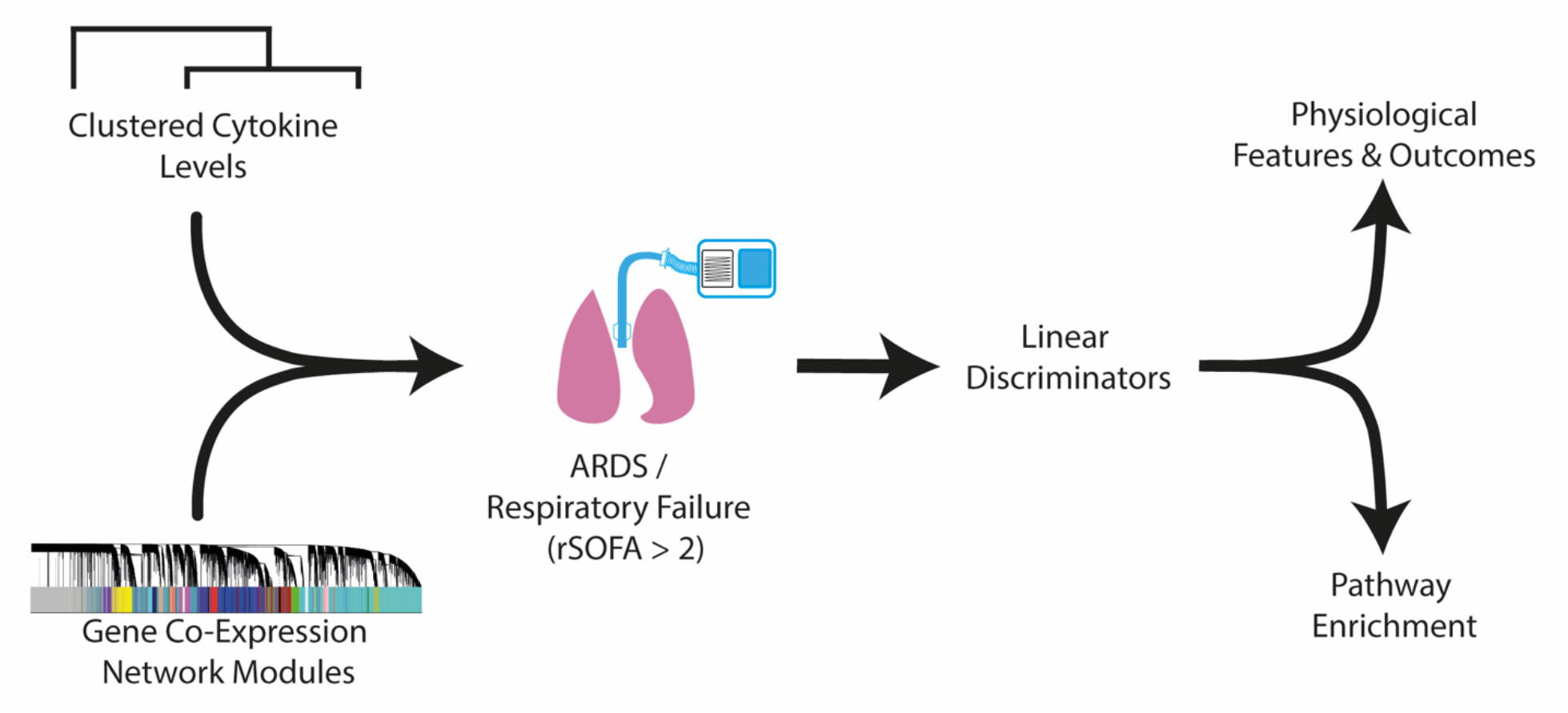
Schematic of the data analysis methods for patients recruited to the MOSAIC study. Measured plasma cytokines were scaled on the mean, and clustered using agglomerative, hierarchical Ward clustering with average linkage. The optimum cluster number was confirmed using the silhouette method and Dunn index. Three clusters had the best fit. Gene expression modules were identified by constructing an adjacency matrix of highly correlated co-expressed genes using a weighted gene network analysis method (WGCNA). This identified genes that were consistently co-expressed in the network and clustered them into gene expression modules known as “module eigengenes”. The modules eigengenes represented the first principal component of the clustered genes within the module. We stratified patients by their respiratory SOFA (sequential organ failure assessment) score to identify the combination of cytokine clusters and gene expression modules that differentiated the most unwell patients between each cluster. In order to determine the variables that were most discriminant between the identified groups, a linear discriminator model was used. This method seeks to maximize the variance between assigned groups in the principal component space. Key gene expression modules underwent pathway and enrichment analysis (www.metascape.org) to identify the mechanisms in each of the patient groups.

**Figure 2:**
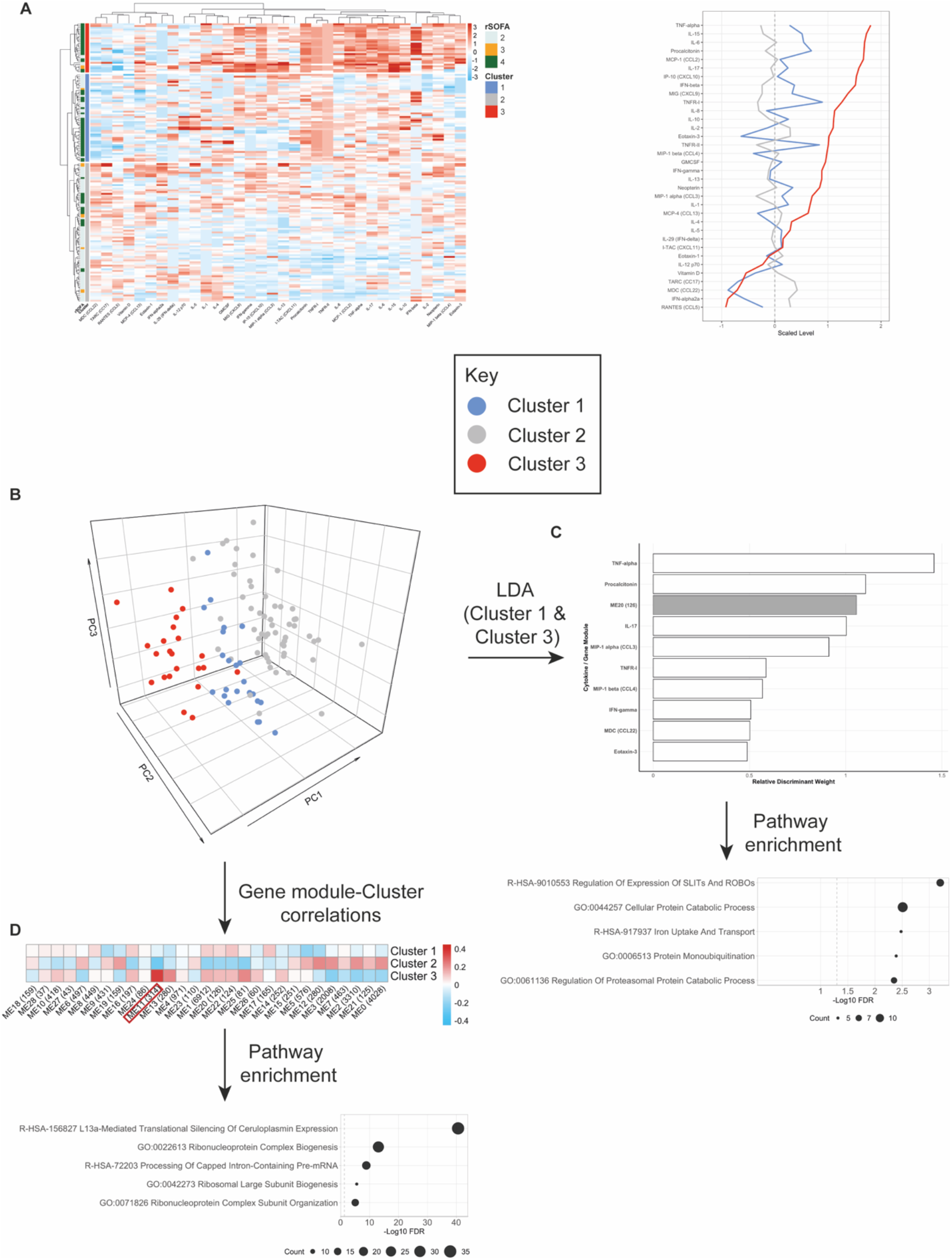
Hierarchical clustering of patient cytokine and chemokine levels was used to determine sub-types of immune response to influenza infection. Identifying key gene expression modules that differentiated patients between clusters was used to determine key mechanisms. **A** Heatmap showing the relative cytokine levels in each cluster from the MOSAIC study. The column on the left side of the heatmap shows the severity of respiratory failure as determined by the respiratory SOFA (rSOFA) score. A large proportion of patients in cluster 1 (blue) and 3 (red) had an rSOFA > 2. The right panel showed the scaled cytokine levels in each cluster, ranked for the highest levels in cluster 3 (red). This cluster appears to be dominated by cytokines and chemokines associated with innate immunity and neutrophil activation (TNF-α, IL-6, procalcitonin). **B** Principal component plot of combined cytokine and module eigengene eigenvalues. Weighted gene expression network analysis (WGCNA) was used to construct a matrix of highly co-expressed genes which were clustered into modules known as module eigengenes (ME). This represents the first principal component of gene expression modules. Eigenvalues for each module with respect were combined with the relative cytokine levels for each patient and plotted in principal component space. Each patient is colored by the cluster membership assigned to it based on the hierarchical clustering of scaled cytokine levels. **C** Linear discriminant analysis of the combined module eigengene and cytokine levels with respect to patients assigned to the blue and red clusters who had severe respiratory failure (rSOFA > 2). The results of this method produced a coefficient matrix where the relative weights of the contributing covariates given a scaled value. This is referred to as the relative discriminant weight. Variables with positive coefficients are strongly differentiating between the assigned group labels, whereas variables with negative coefficients do not differentiate data points between the assigned groups. Of the top 10 discriminant elements, only one related to a gene module. The top pathway associated with this module enriched for genes was associated with SLIT-ROBO expression. **D** Correlation matrix (Pearson’s) between each of the clusters and gene expression modules. Only one module was significantly differentially correlated between clusters: ME11 which contained 314 genes. The top pathway enriched for this gene module was ‘L13a mediated silencing of caeruloplasmin’. The relates to an IFN-γ induced pathway which suppresses the expression of genes involved in the acute phase of the inflammatory response. This is known as the gamma-interferon associated inhibition of transcription (GAIT) mechanism. Patients in cluster 3 (red) had the highest levels of IFN-γ, whilst patients in cluster 2 (grey) had the lowest levels (also see panel **A**). LDA linear discriminant analysis FDR false discovery rate ME module eigengene

### Network analysis combined with linear discrimination identifies the most important contributing modules when comparing clusters

Weighted gene co-expression network analysis (WGCNA) identified 29 gene modules in the MOSIAC cohort. Of these modules, one consisting of 314 genes, was positively correlated with cluster 3 (Pearson’s correlation coefficient +0.36, p = 0.005). The same module was negatively correlated with cluster 2 (Pearson’s correlation coefficient −0.27, p = 0.16) (Figure 2D).

The module eigengene values were combined with the scaled biomarker concentrations. Principal component projection of these data showed the eigengene values to be orthogonal to the biomarker data. A linear discriminant method (LDA) ranked the relative weights of biomarkers and eigengenes, using the cluster labels as a supervised classifier. Linear discrimination between clusters 1 and 3 found TNFα, procalcitonin, and IL-17 to be the most important factors differentiating them (Figure 2B); This was consistent with the averaged scaled concentrations of these biomarkers (Figure 2A). Only one gene module (containing 126 genes) had a high variable importance when using this method (Figure 2C). In order to determine the most important variables for each of the clusters, we used the multi-classifier property of LDA, which can find the relative importance of contributing variables in the context of more than two supervised labels. The model AUROC was equal to 0.88 using this method.

### Pathway enrichment of key gene modules reveals mechanistic insights

Cluster 1 was differentiated from both clusters 2 and 3 by the same 126 gene module. Gene set enrichment analysis (ww.metascape.org) showed a significant over-representation for SLIT-ROBO signaling pathway (R-HSA-90110553) after adjustment for multiple comparisons (q = 3.2). SLIT-ROBO signaling is recognized to play a role in neuronal axonal development but has more recently been shown to be important in pulmonary endothelial barrier integrity in murine models of sepsis and influenza. Slit2-Robo4 receptor interactions preserved murine pulmonary endothelial V-cadherin levels despite local inflammatory stimuli *(13)*. The same mechanism has also been shown to be involved in endothelial leak using *in vitro* human endothelial cell models of transfusion-associated lung injury (TRALI) and hantavirus infection *(14, 15)*.

Cluster 3 was strongly positively correlated with a gene module containing 314 genes. The same module was negatively correlated with cluster 2. The genes from this module showed a significant over-representation for a pathway entitled ‘L13a-mediated translation silencing of caeruloplasmin’ (R-HSA-156827, q = 40.6). This process is related to the gamma-interferon inhibition of translation (GAIT) mechanism where, under the influence of IFN-γ, ribosomal proteins are able to bind 3’ mRNA motifs (GAIT motifs) and prevent their translation *(16)*. Caeruloplasmin is an acute phase protein, and this mechanism is thought to be a late response to IFN-γ that helps to ameliorate innate immune responses by inhibiting translation of these mRNA.

### Mechanisms are consistent with clinical and immunological features of these patients

Patients in cluster 3 and 1 were significantly more likely to have received mechanical ventilation, and to have significantly lower serum albumin concentrations (Figures 2 and 3). Patients in cluster 3 also had evidence of multi-organ dysfunction, with significantly higher bilirubin and creatinine concentrations than the other clusters (Figure S2). A large proportion of patients in cluster 1 had a requirement for mechanical ventilation and very low albumin, but had no evidence of multi-organ failure, thus we described these patients as being characterized by isolated endothelial leak.

**Figure 3:**
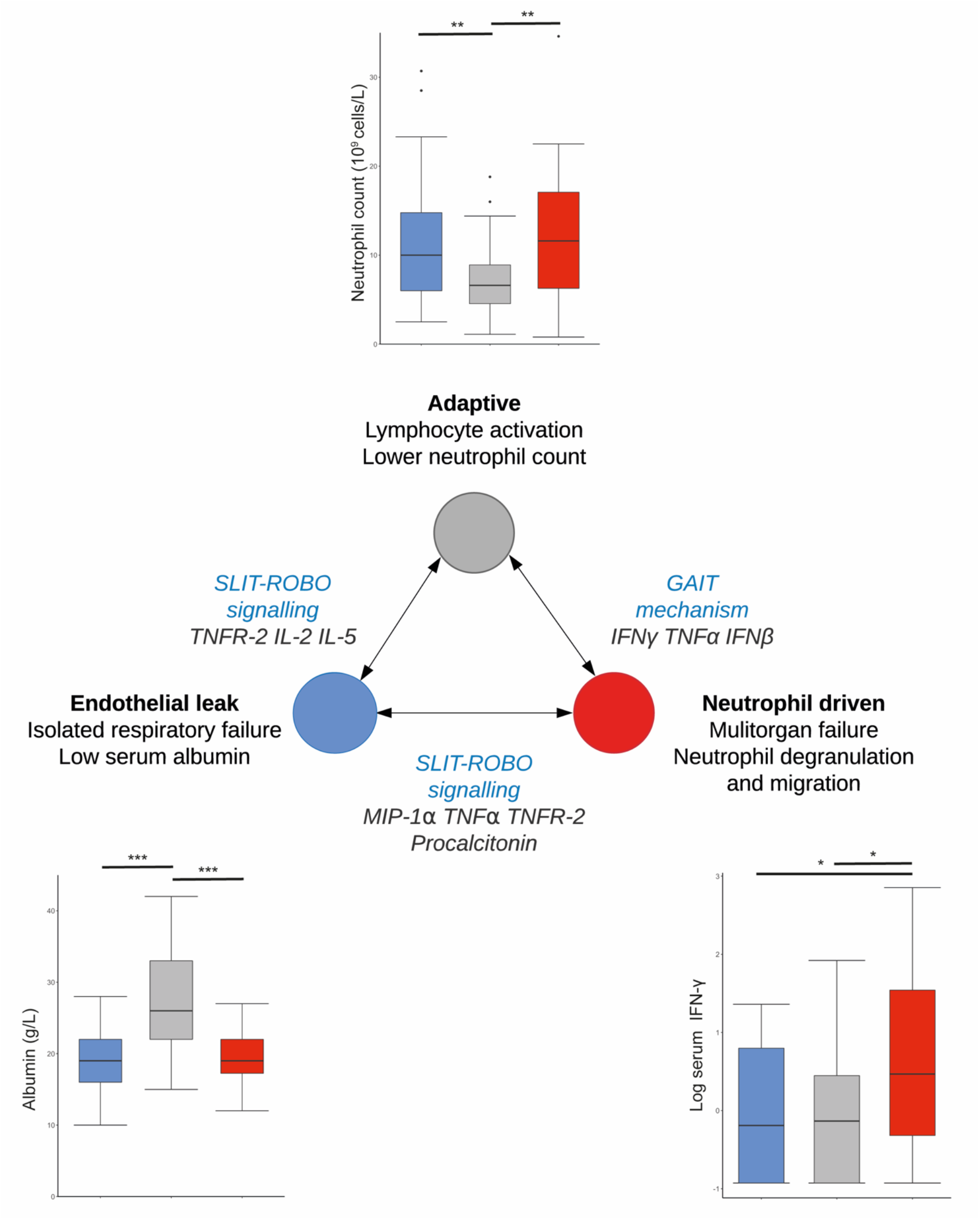
Summary of the key cytokines, chemokines and gene modules that differentiated patients between each endotype as determined by linear discriminant analysis and Pearson’s correlation coefficients. The peripheral boxplots show some of the clinical variables that differentiated each of the endotypes. Patients with the endothelial leak endotype were delineated from the other endotypes by a gene module that was enriched for SLIT-ROBO signaling; a mechanism that preserves endothelial barrier function. These patients had a high rate of respiratory (but not multiple) organ failure and low albumin levels. Patients with the neutrophil-driven endotype were characterized by increased expression of genes that were enriched for pathways relating to neutrophil activation and degranulation. These patients also had significantly higher levels of serum IFN-γ which was consistent with finding that the key differentiating gene module was enriched for L13a mediated translational silencing of caeruloplasmin expression pathway. Patients with this endotype also had significantly higher creatinine and bilirubin. Patients in the adaptive endotype showed low levels of TNFα, IL-6 and TNFR1 and TNFR2, had significantly higher serum albumin concentration, lower peripheral blood neutrophil counts, were less likely to have an rSOFA of 3 or 4, and had higher expression of genes that enriched for pathways associated with lymphocyte activation and adaptive immunity. TNF tumor necrosis factor, TNFR tumor necrosis factor receptor, IFN interferon, MIP-1α macrophage inhibitory protein (CCL3), GAIT gamma interferon associated inhibition of transcription. * p < 0.05, ** p < 0.01, *** p < 0.001. All p-values were corrected for multiple comparisons.

Patients in cluster 3 had significantly higher concentrations of acute phase biomarkers (TNFα, IL-6, GM-CSF, IL-8) and cytokines such as IL-17 that induce release of these markers (Figure 2). These patients also had high concentrations of cytokines associated with anti-viral immune responses (IFN-γ, IL-15) (Figures 2 and 3). Despite the substantially increased concentrations of IFN-γ and activation of genes associated with the GAIT mechanism, this process was likely failing to suppress the acute phase of illness, suggesting failure of the immune mechanisms required to down-regulate an unwanted innate immune response.

### Differential gene expression reveals additional mechanistic insights into the identified clusters

Differential gene expression between patients with severe respiratory failure in clusters 3 and 2 identified 514 genes that were significant (FDR < 0.05, Figure S3). Pathway enrichment for significantly upregulated genes (116) in cluster 3 showed the ‘neutrophil degranulation’ pathway (GO:0043312, q = 8.9) to be significantly over-represented. Twelve genes had a fold change greater than 1.5, of which CD177, MMP-8 and OLM4, RETN have all previously been associated with neutrophil activation in severe influenza *(17)* and ARDS *(18, 19)*. ZDHHC19 has also been associated with increased expression in patients with ARDS *(18, 20, 21)* and sepsis *(22)*, across different microarray platforms. Neutrophil counts were significantly increased in patients within cluster 3 compared to those within cluster 2, but were similar to those with the endothelial leak endotype (cluster 1). We describe this endotype ‘neutrophil-driven’.

The 398 genes that were significantly upregulated in cluster 2, compared with the neutrophil-driven endotype, enriched for pathways relating to ‘lymphocyte activation’ (GO:0046649, q = 11.1) and ‘adaptive immunity’ (GO:002250, q = 7.4, Figure S3). When compared with the endothelial endotypes, pathways enriched for ‘hemostasis’ (R-HAS-109582, q = 17.66) and ‘regulation of wound healing’ (GO:0061941, q = 4.83) were over-represented (Figure S3). We labelled this endotype ‘adaptive’ due to its significantly lower concentration of immune mediators associated with the innate response, raised concentrations of chemokines associated with T cell function (RANTES, TARC, MDC) and association with adaptive immunity pathways (Figures 2A and S3).

There were only four differentially expressed genes between the neutrophil-driven and endothelial leak endotypes. None of these enriched for a pathway, highlighting that genomics alone is unable to discriminate between these groups of patients.

### Endotypes are stable at 48 hours and associated with different outcome profiles

A repeat set of patient sampling for biomarkers was taken on the second day (T2) after recruitment in 53 of the 104 patients; This was taken at median interval of 2.2 days after the first sample. The transitions between clusters over time is shown in figure 3. Where data were available, clusters were stable with little transitioning between them. 18.5% of patients with the adaptive endotype were discharged home by T2. Although a high proportion of patients with the endothelial leak endotype required mechanical ventilation (48.5%), their 30-day survival was comparable to the adaptive group (HR=0.79, 95%CI 0.11 – 5.72). The neutrophil driven endotype had a significantly worse 30-day survival (HR=5.15, 95%CI 1.04 – 25.6) (Figure 5).

**Figure 4:**
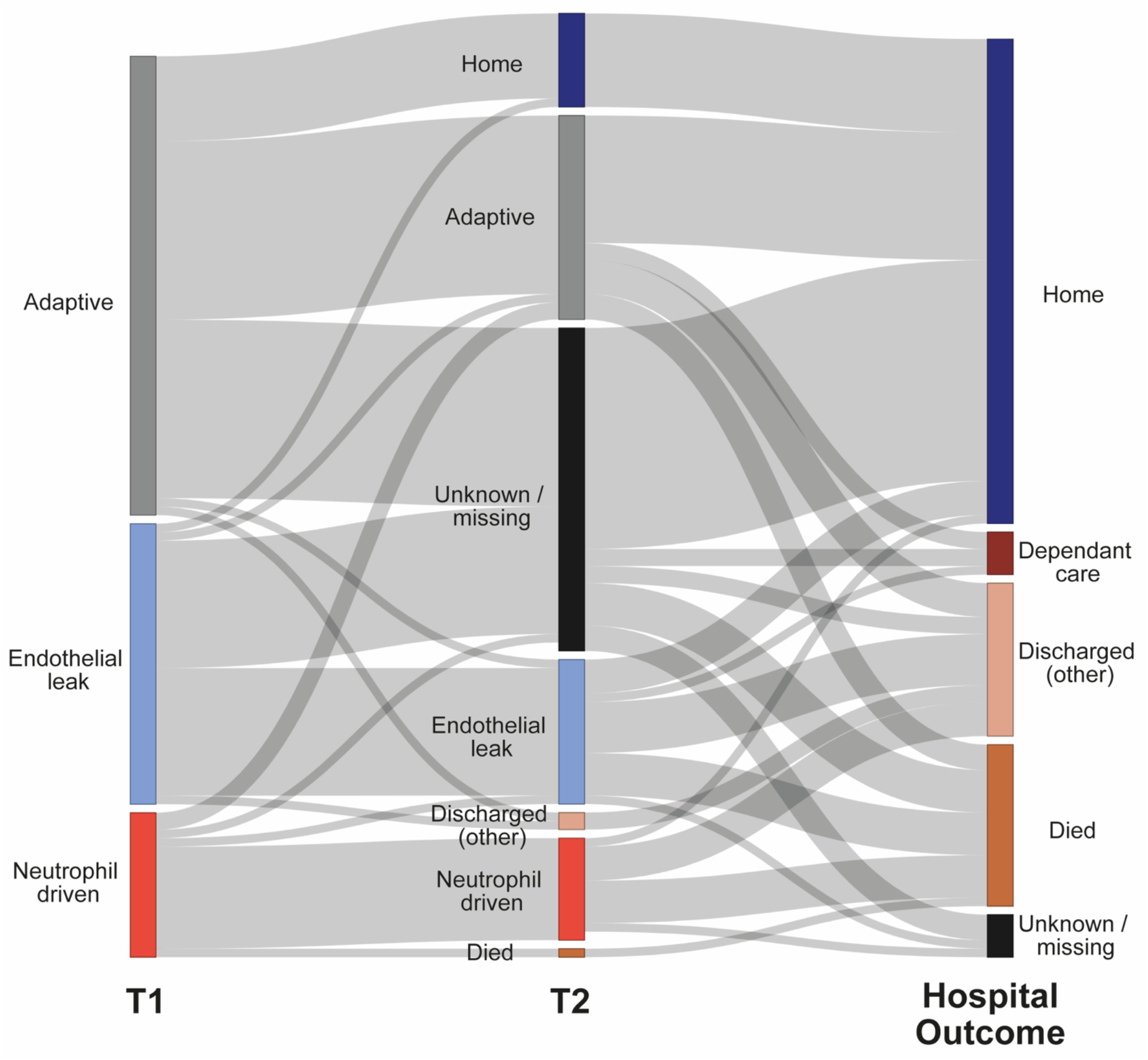
Stability of endotypes between admission to hospital (T1), 48 hours after admission to hospital (T2), and hospital discharge. The median sampling time at T2 was 2.2 days. Where data were available, there was very little transitioning between endotypes over this time period. Given that that there would have been variation between patients presenting to hospital in terms of their symptom duration and disease severity, endotypes must be stable for our methods to have detected them in this fashion. ‘Unknown / missing’ refers to patients who did not undergo sampling or were lost to follow up. ‘Dependent care’ refers to discharge to residential, supported living or with relatives. ‘Discharged (other)’ refers to other discharge locations, which were community or local hospitals.

**Figure 5.**
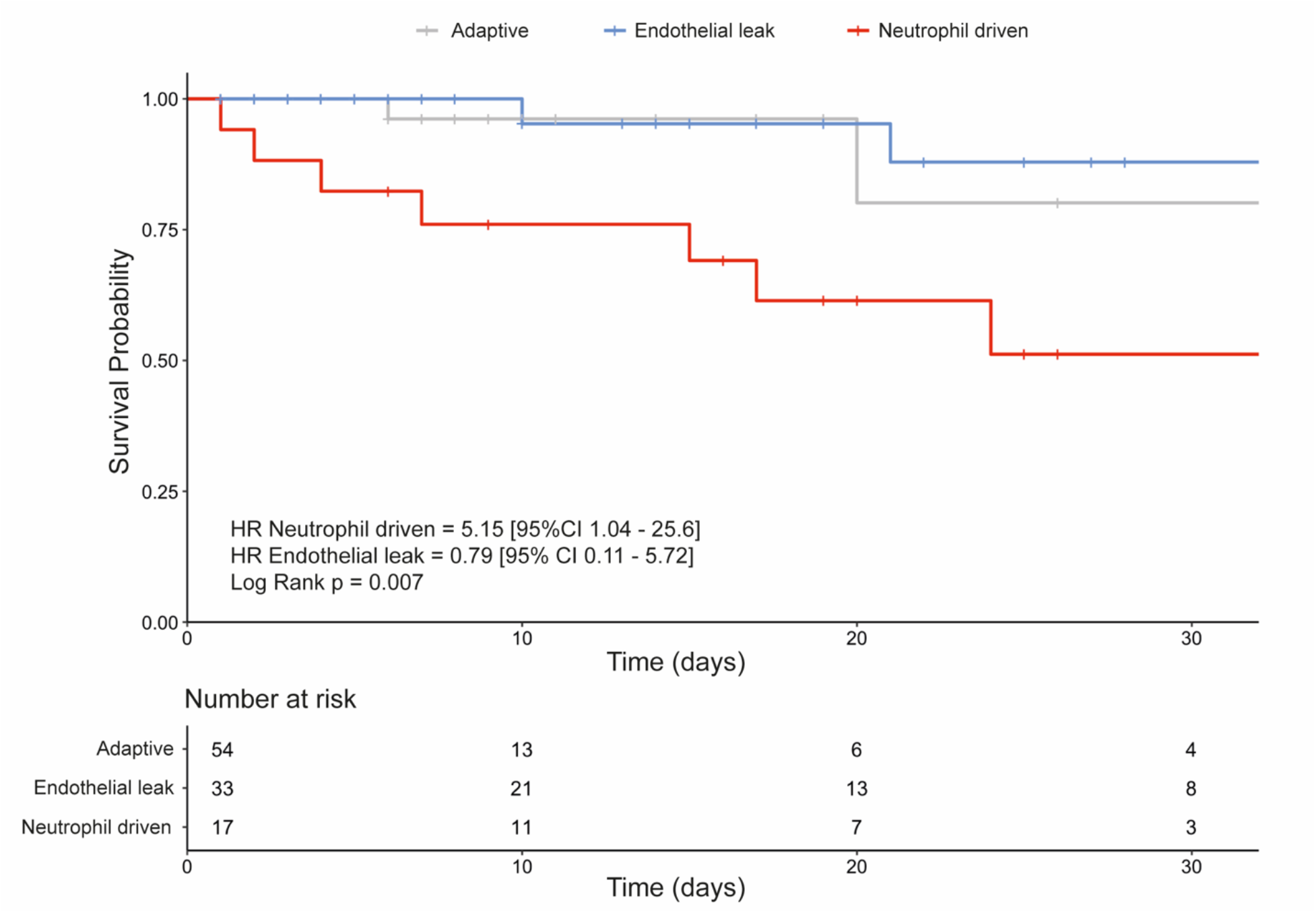
Kaplan-Meier curve showing 30-day survival of patients assigned to each endotype at T1. Hazard ratios were calculated using Cox Proportional Hazard regression. The adaptive endotype was the reference group for outcome comparisons. Crosses represent censoring.

Patients with the adaptive (88.9%) or endothelial leak endotypes (69.7%) were more likely to be discharged from hospital at 30 days than the neutrophil-driven endotype (41.1%); OR Adaptive = 11.4 [3.29-44.4, p = 0.0002], OR Endothelial leak = 3.29 [95%CI 0.99-11.6, p = 0.055]).

### Biomarker sampling from the posterior and anterior nasopharynx does not mirror serum levels

77 patients with rSOFA ≥ 2 had biomarkers sampled simultaneously from their serum, anterior nasopharynx (SAM), and posterior nasopharynx (NPA). Figures 6 shows the relative concentrations in each body compartment for each of the described endotypes, and that they poorly correlated with each other. Hierarchical ward clustering on the nasal immune mediators revealed three clusters in each of the anterior and posterior nasal compartments. The clusters poorly correlated with the serum clusters (Figure S4). One cluster from the NPA was strongly associated with rSOFA of 4, with increased concentrations of IL-6, IL-8 and IL12-p70.

**Figure 6:**
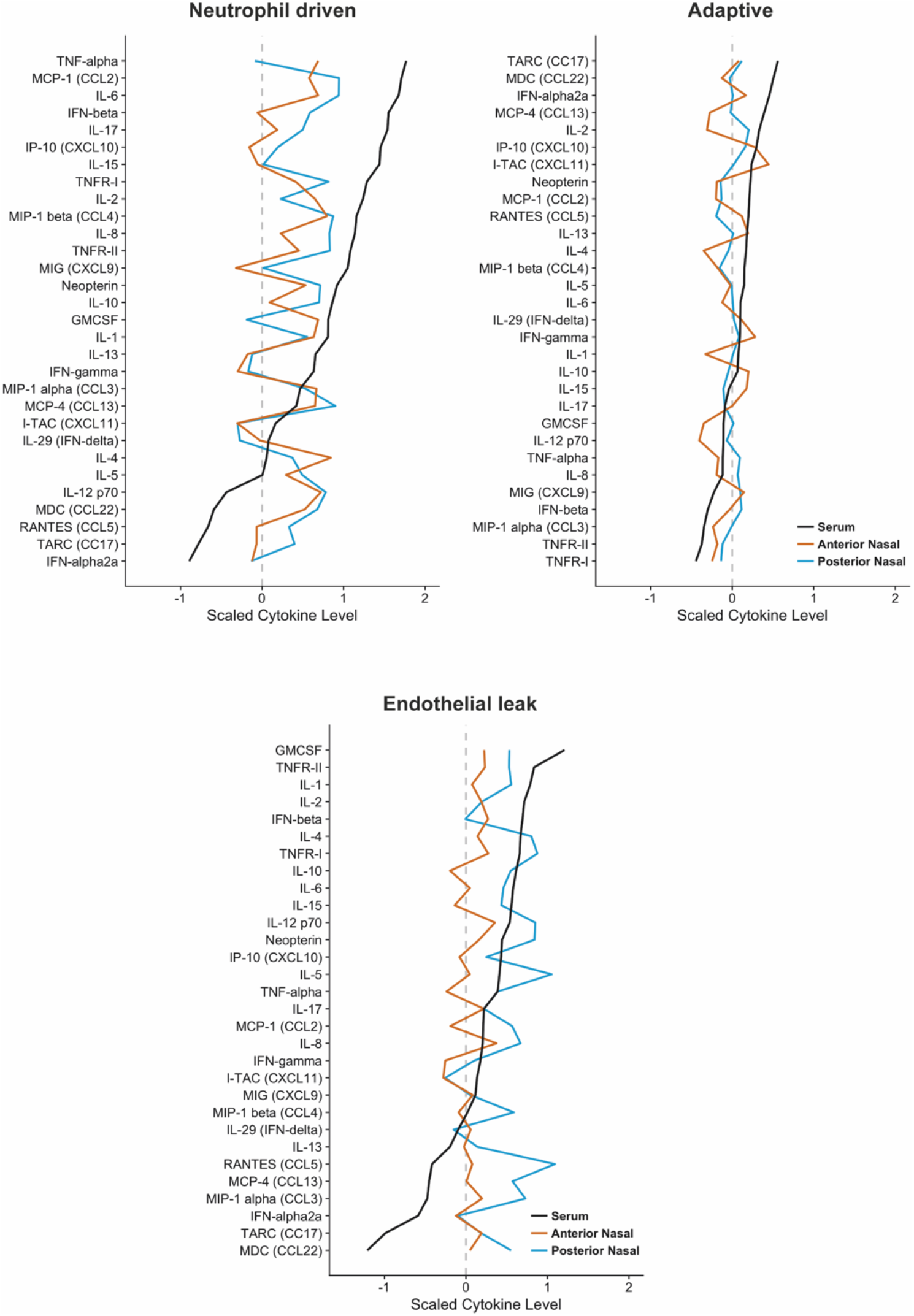
Comparison of averaged, relative cytokine levels in serum, anterior nasal and posterior nasal compartments for patients in the neutrophil driven and adaptive endotypes. 77 patients had their soluble immune mediators measured in all three compartments. Measured cytokines were log-normalized for each compartment before averaging across all patients in each endotype. Measured nasal cytokines poorly reflected the serum levels of the same cytokine or chemokine for patients in each endotype. Relative cytokine levels are represented on the same z-score scale across all three endotypes.

### Bacterial co-infection does not account for the differences between endotypes

Extensive screening for bacterial infection took place as part of the MOSAIC study, including culture and PCR of both nasopharyngeal and sputum samples, urinary pneumococcal antigen testing, and review of the results of the microbiological cultures ordered by the treating clinical teams. An expert panel then determined if positive results relating to pathogenic bacteria were clinically relevant. Of the 104 patients with serum biomarker and gene expression data, 44 (42.3%) were deemed to have been adequately screened for secondary bacterial infection using at least four of the above modalities. We only considered T1 endotype assignments as there was no way of knowing whether patients had been successfully treated for bacterial infections by T2. In total, 25 patients (56.8%) were deemed positive for clinically significant bacterial infection. 83.3% of successfully screened patients with the neutrophil-driven endotype had clinically relevant bacterial co-infection, but the infection rates were also high in the other two endotypes (adaptive: 56%, endothelial leak 57%) and this difference was not statistically significant (OR 3.9, 95%CI 0.82-29.1, p = 0.12) (Table 2). A higher proportion of the patients in the neutrophil driven endotype were successfully screened for infection (71%) than the other endotypes (adaptive: 46%, endothelial leak: 21%), this might be reflective of the increased proportion of patients with this endotype being managed in an intensive care, where invasive sampling is often part of routine clinical care. Bacterial co-infection thus may contribute to a neutrophil-driven response in influenza, but this alone does not define an endotype given the evidence of a high proportion of bacterial co-infection in the other two groups.

**Table 2:** Patient outcomes by endotype. Note data here is limited to patients who had both gene expression and immune mediators measured (n = 104). Categorical variables were tested by logistic regression. The difference between endotypes for each is expressed by their odd ratios (OR) and their 95% confidence limits with respect to the hypo-inflammatory endotype which was used as the reference category. The length of stay data was log transformed prior to fitting a Poisson regression model. Odds ratios cannot be easily interpreted for these types of models and so only the p-values of the model coefficient estimates are provided for this variable.

|  | **Adaptive** | **Endothelial leak** | **Neutrophil driven** |
| --- | --- | --- | --- |
| Adequately screened for evidence of bacterial infection*, n (%) | 25<br>(46%) | 7<br>(21%) | 12<br>(71%) |
| Clinical evidence of bacterial infection, n (%) | 11<br>(56%) | 4<br>(57%) | 10<br>(83.3%) |
|  | *Reference* | *OR 0.59*<br>*(0.1-3.2)*<br>*P = 0.54* | *OR 3.93*<br>*(0.82 – 29.1)*<br>*P = 0.12* |
| Admitted to critical care, n (%)† | 13<br>(24.1%) | 20<br>(60.6%) | 14<br>(82.4%) |
|  | *Reference* | *OR 4.85*<br>*(1.94 – 13.7)*<br>*P < 0.001* | *OR 14.7*<br>*(4.07 – 71.6)*<br>*P < 0.001* |
| Mechanical ventilation, n (%)† | 11<br>(20.4%) | 16<br>(48.5%) | 11<br>(64.7%) |
|  | *Reference* | *OR 3.68*<br>*(1.44-9.8)*<br>*P = 0.007* | *OR 7.2*<br>*(2.25-25.2)*<br>*P = 0.001* |
| Vasopressor support, n (%)† | 8<br>(14.8%) | 17<br>(51.5%) | 10<br>(58.8%) |
|  | *Reference* | *OR 6.11*<br>*(2.3-17.6)*<br>*P < 0.001* | *OR 8.2*<br>*(2.49-29.5)*<br>*P < 0.001* |
| Median length of hospital stay, days, [IQR] | 6<br>[2.25 – 9.75] | 15<br>[4-26] | 20<br>[9.5-30.5] |
|  | *Reference* | *P = 0.001* | *P = 0.04* |
| Hospital Mortality, n (%) | 4<br>(7%) | 7<br>(21%) | 8<br>(47%) |
|  | *Reference* | *OR 3.36*<br>*(0.93 – 13.8)*<br>*P = 0.07* | *OR 11.1*<br>*(2.90 – 49.6)*<br>*P = 0.007* |
| 30-day mortality, n (%)‡ | 2<br>(3.7%) | 2<br>(6.45%) | 6<br>(35.3%) |
|  | *Reference* | *OR 1.67*<br>*(0.19 – 146)*<br>*p = 0.61* | *OR 14.2*<br>*(2.85 – 106.2)*<br>*p = 0.003* |

\* at least four modalities of NPA (culture and PCR), sputum (culture and PCR), pneumococcal urinary antigen, blood cultures (if taken by treating clinical team)
† during the first 14 days after recruitment.
‡ based on in-hospital mortality only (out of hospital data not available).
OR: Odds ratio

## Discussion

Here we present the outcome of a novel bioinformatic analysis of patients with moderate/severe influenza infection and identify three distinct patient endotypes with underlying plausible biological mechanisms, providing new insights into host responses to acute influenza infection.

The three endotypes identified can be described as “neutrophil-driven”, “adaptive”, and “endothelial leak”. The neutrophil driven patients display evidence of innate immune activation (high levels of TNFα, IL-6, IL-8, IL-17) with associated multi-organ dysfunction and reduced 30-day survival. These patients could be differentiated from the adaptive endotype by an alteration in the GAIT-mechanism, a late transcriptional regulatory response to IFN-γ that acts to suppress innate immunity by reducing caeruloplasmin mRNA translation *(16)*. Patients with the neutrophil-driven endotype had significantly increased IFN-γ levels but appeared unable to suppress their innate immune response (figure 2a). The endothelial leak endotype could be distinguished from both the neutrophil driven and the adaptive endotypes by alterations in Slit-Robo signalling, a pathway important in the maintenance of endothelial barrier integrity, and although patients in this group required mechanical ventilation, they did not develop multu-organ failure in the manner of the neutrophil-driven endotype patients.

The mechanisms that underlie our three endotypes can be explained by exploring the immunopathogenesis of influenza. The influenza A non-structural 1 protein (NS1) suppresses IFN-γ signaling by preventing STAT1-mediated phosphorylation of cyclin-dependent kinase 5 (CDK5) *(23)*. CDK5 and CDK5-dependent AGC kinase are responsible for phosphorylation of glutamyl-prolyl tRNA synthetase (EPRS) *(16)*. Dual phosphorylation of EPRS releases it from the multi-synthase complex allowing it to form the pre-GAIT complex with eIF3. EPRS release is the first stage of the GAIT mechanism and occurs 2-4 hours following IFNγ stimulation. The second stage of the GAIT mechanism involves phosphorylation of the ribosomal protein, L13a, about 16 hours after IFNγ stimulation. L13a is released from the 60s ribosome and combines with GAPDH and the pre-GAIT complex to form the GAIT complex. The GAIT complex is able to bind hairpin loop-like motifs in the 3’ untranslated regions of mRNA, and prevent translation of the bound RNA molecules *(24)*. EPRS also plays an anti-RNA viral role by binding poly(rC) binding protein 2 (PCBP2). PCBP2 inhibits the ubiquitination of the mitochondrial antiviral system (MAVS) of proteins. MAVS, via TRAF4, induces the transcription of type 1 interferons *(25)*. It has also been noted that the coronavirus SARS-CoV interferes with this mechanism as its ORF9b protein promotes degradation of both MAVS and TRAF4 *(26)*. Thus, virally induced alterations in the GAIT mechanism provide a plausible mechanism via which patients displaying the neutrophil driven develop multiorgan failure and dysregulated innate immune activation.

Interferon-related mechanisms have been shown to be of importance in non-influenza viral infections. The alpha coronavirus transmissible gastroenteritis (TGEV), has been observed to contain a GAIT motif within its genome that is able to arrest translation, and exacerbate the host innate immune response *(27)*. TGEV is distinct from the well-known epidemic/pandemic beta coronaviruses SARS-CoV, MERS, and SARS-CoV2. Clinical data from critically ill patients with SARS-CoV also identified a key role for IFN-γ in severe disease *(28)*. Disruption of IFN-γ related mechanisms by viral proteins may directly drive pathogenesis since IFNGR1 knockout mice show reduced pathological changes following influenza A challenge *(29)*, suggesting that RNA viruses may have evolved mechanisms to disrupt the immunomodulatory effects of IFN-γ, which has roles in both innate immune regulation and anti-viral gene expression.

Differential gene expression between endotypes revealed upregulated genes associated with neutrophil activation, transmigration and degranulation in both the neutrophil-driven and endothelial leak endotypes. (Figure S3). Of note we found increased expression of CD177, which encodes a glycosyl-phosphatidylinositol (GPI)-linked cell surface glycoprotein that has recently been implicated in acute influenza infection *(17)*. CD177 is expressed on the cell surface of neutrophils and is thought to play a role in cell-cell interactions via beta-2-integrins and platelet endothelial cell adhesion molecule 1 (PECAM-1). CD177 is usually surface bound and cannot be detected in serum. Two other genes with increased expression in the neutrophil-driven and endothelial leak endotypes, ZDHCC19 and RETN, have also been reported to be upregulated in peripheral whole blood and bronchoalveolar neutrophils obtained from patients with ARDS *(18, 20)*. Each of these studies used different microarray platforms, with different probe configurations, suggesting that our findings of upregulated genes associated with neutrophil activation, transmigration and degranulation in the neutrophil driven endotype are robust, given that they are consistent with these independent analyses.

A significant proportion of the patients with the endothelial leak endotype required mechanical ventilation. These patients had low serum albumin, and despite having a clinical picture suggestive of poor outcome, had similar 30-day survival to the adaptive endotype and significantly better 30-day survival than the neutrophil driven endotype (Figure 4). The endothelial leak endotype was characterized by gene modules enriched for the expression of the SLIT-ROBO pathway. Slit2-Robo4 receptor interactions are responsible for maintaining the endothelial barrier by preserving V-cadherin expression at the cell surface in murine models of influenza infection *(13)*. Slit-2 can abrogate the pulmonary endothelial permeability induced by influenza infection, and leak in human endothelial cell models of hantavirus infection and transfusion-associated lung injury *(13–15)*.

Patients in the adaptive endotype had reduced concentrations of innate immune mediators and increased concentrations of mediators associated with T cell activity. Differential gene expression identified pathways associated with the adaptive immune response, haemostasis and wound healing. Although the differentially expressed genes also enriched for the lymphocyte activation pathway, the lymphocyte counts in these patients were not significantly different to those in other endotypes highlighting the limitations of standard clinical tests for identifying disease subtypes. Patients displaying the adaptive endotype were most likely to have returned home by T2, compared with the other two endotypes.

The stability of endotypes over the 48-hour sampling interval, and the improved survival of the endothelial leak endotype in comparison with patients in the neutrophil driven endotype, suggests that the three endotypes are distinct entities. Their persistence over this time period also suggests that they may be amenable to future study, and importantly, to being used to stratify patients for enrolment into clinical trials.

We were unable to demonstrate the same endotypes when using immune mediators in the posterior (NPA) or anterior (SAM) compartments of the nasopharynx (Figure 6). Scaled immune mediator levels in the neutrophil-driven endotype patients had similar patterns for the anterior and posterior nasopharynx, but these did not reflect the concentrations of the same mediators in the serum. Hierarchical clustering and heatmap analysis revealed three optimal clusters in both the SAM and NPA samples, however, these did not align with serum clusters (figure S4). In the NPA samples there were three identified clusters. Patients in the cluster with high levels of innate immune mediators (IL6, IL8, IL12p70) all had an rSOFA score of 4. In the cluster with globally depressed immune mediators, 34% received mechanical ventilation. These two clusters did not align with the neutrophil driven or adaptive endotypes. Immune mediators from the SAM samples neither aligned with rSOFA, nor serum based endotypes, suggesting that the nasal pharyngeal compartment poorly reflects the signals found in the blood compartment. The larger mass of lung tissue and the flow of the entire blood volume through the lungs approximately once a minute might explain the close alignment of the lung and blood signal.

Hyperinflammatory phenomena have been described previously in influenza infection and are known to be associated with poor outcomes *(30, 31)*. However, our analyses demonstrate that there are three distinct mechanistic processes occurring in patients with moderate or severe influenza, which relate to differential clinical outcomes. Given that a large proportion of patients in the endothelial leak endotype required mechanical ventilation and vasopressor support, it is surprising that their 30-day mortality was similar to those with an adaptive immune response to influenza. The lack of differentially expressed genes between the endothelial leak and neutrophil driven endotypes is particularly striking given their different immune mediator profiles and clinical features. This brings into question the value of comparing individually expressed genes in isolation, when trying to explain immunological and clinical phenomena in complex multi-system diseases. Given that genes work in coordinated networks, the WGCNA approach we adopted allowed us to identify gene modules responsible for differentiating biological processes between these subgroups of patients.

The combination of a biomarker panel with standard testing early in a patient’s admission offers a novel approach to early patient stratification, enabling differential clinical management approaches and structured inclusion into future randomized controlled trials. For example, it is possible that endothelial leak patients may not need mechanical ventilation if interventions targeted at preserving/restoring endothelial function can be adopted early. On the other hand, for patients with the neutrophil-driven endotype, specific immunomodulation of the host response may be effective in altering the clinical outcome. Patients found to be in the adaptive group at recruitment to a future trial might not require intervention at all, given their better outcomes; This would reduce the risk of exposure of these patients to adverse events from an investigational intervention, potentially worsening their outcomes and reducing the relative effect size of any benefit from the intervention in other study arms. Future work should focus on validating these endotypes prospectively in the context of viral infection, to refine our understanding of the underlying mechanisms in order to develop tools to allow the prospective stratification of patients for targeted therapeutic interventions.

## Materials and Methods

### Study design and participants

The mechanisms of acute severe influenza consortium (MOSAIC) prospectively recruited adult patients admitted to participating UK hospitals with suspected influenza infection during the pandemic influenza A seasons of 2009/10 and 2010/11. Patients with confirmed influenza infection underwent biological sampling at three time points: T1 (recruitment), T2 (2 days after recruitment) and T3 (at least four weeks after T1).

This study sought to identify the mechanisms in patients with severe respiratory failure following influenza infection, and if these were also common to patients who developed ARDS due to community acquired pneumonia.

### Ethical approvals

The MOSAIC study was approved by the NHS Research ethics service, Outer West London REC (09/H0707/52, 09/MRE00/67). Patients or their legally authorized representatives provided consent for recruitment to both of these studies.

### Immune profiling and viral infection status

The methods by which samples were collected and processed by the MOSAIC study consortium are described previously *(11)*. 34 soluble immune mediators were measured in the blood of recruited patients. 30 of these were measured using multiplex electrochemiluminescence methods on the MSD SECTOR instrument (Mesoscale Discovery, Gaithersburg, USA). RANTES (CCL5), neopterin and Vitamin D were measured using ELISA and plasma procalcitonin using the Elecsys BRAHMS PCT assay on a calibrated Cobas e602 platform.

The posterior nasopharynx was sampled using a 10Fr Argyle catheter inserted into the nostril which was instilled with 10 mL sterile saline and gentle suction applied. The anterior nasopharynx was sampled using a synthetic absorptive matrix (SAM, LeukoSorb Pall) which was applied to the lateral nasal wall via the nares. This fibrous matrix binds mediators in the fluid adsorbed onto the strips which can later be separated from the by elution-centrifugation. 31 soluble immune mediators were measured in the anterior and posterior nasopharynx using the same multiplex electrochemiluminescence methods on the MSD SECTOR instrument.

### Gene expression profiling

The MOSAIC study collected 3 mL of whole blood into two Tempus tubes (Applied Biosystems, Ambion), stored at −80°C for RNA extraction. RNA extraction methods for are described elsewhere *(11)*. Illumina Human HT12 V4 Expression BeadChips microarrays with 47,231 probes (Illumina, San Diego, CA, USA) were used to do genome-wide transcription profiling on purified RNA samples. Microarray results were log transformed, background corrected and ‘rsn’ normalized using the *lumi* R package. Probes with low expression levels were removed using their relative detection calls.

### Bacterial Infection Status

Samples collected from the nasopharynx and throats of patients by the MOSAIC study investigators were cultured for bacteria and underwent multiplex PCR for common respiratory organisms. Where available, urine samples were tested for pneumococcal antigen (BinaxNow, Alere). Microbiological culture results from tests initiated by clinical teams were also collected and stored in the database. A panel of clinical experts independently assessed these results to determine the bacterial infection status of recruited patients and exclude positive samples contaminated by commensal organisms.

### Bioinformatics and statistical analysis

Microarray data were variance stabilization normalized. Weighted gene co-expression network analysis (WGCNA) is a method of identifying groups of genes that are consistently expressed or co-regulated *(8, 32)*. A network between each pair of genes is constructed based on their relative co-expression across all the measured samples. The similarity between a given set of genes in the network is determined by their relative Pearson’s correlation coefficients. Correlation matrices are then transformed into a scale free adjacency matrix by using a power function (β). The connectivity between genes across this scale free network can then be calculated using a topological overlap. A modified (hybrid) hierarchical clustering method then organizes inter-connected genes into families which are referred to as modules. The first principal component of each module is referred to as its ‘eigengene’. The relationships between each sample and each eigengene is given as a value between −1 and 1. Characteristics of groups of patients (e.g. ARDS) can be correlated with the eigengene values to identify associations between traits and gene modules.

22,816 probes were used as the input for WGCNA following normalization and filtering of the microarray data using the *lumi* package. The soft power threshold was calculated as 7 using the scale free threshold R^2^ > 0.8 method. For this analysis we used a bidirectional (unsigned) network, with deep split equal to 2 and minimum module size of 30 for hybrid clustering.

The immune mediator levels were log transformed and scaled. Normality was checked using quantile-quantile plots. There was no imputation of missing data points. Hierarchical, Ward automated clustering was performed on these data. Ward’s method minimizes the variances between successively identified branches to construct a dendrogram. Optimum cluster number was determined using k-means clustering and within cluster sum of squares (WCSS) calculations to find the curve vertex (the ‘elbow’ method). Consensus clustering indices methods using the *NbClust* R package were also used to confirm the optimum cluster number *(12). NbClust* applies twenty-four indices to clustered data and establishes a consensus opinion by majority vote.

The log scaled immune mediator levels were combined with the eigengene values for each module. Therefore, each patient had a similar number of features from the gene expression and immune mediators. Principal component analysis was used to examine the relationship between patient clusters and this combined data. A linear discriminant method (LDA) was used to identify the gene modules and immune mediators that contributed most to the separation of patients in each cluster. LDA is similar to principal component analysis (PCA) but seeks to maximize the variance between points that have been assigned a label (i.e. supervised). A fitted LDA model assigns relative importance to each contributing covariate. A positive importance means that this feature is strongly discriminant, whereas a feature with a negative importance is not discriminant between assigned groups. LDA can be used to distinguish the discriminant features between multiple (more than two) groups simultaneously, without having to resort to ‘one-vs-all’ based methods that other classification methods use (logistic regression, random forests). Models fits were checked using leave one out cross validation and receiver operating curve statistics (AUROC) with the *multiclass*.*roc* function from the *pROC* R package.

Genes from important modules that discriminated between clusters were submitted for pathway enrichment tools (www.metascape.org) to identify key nodes and canonical pathways *(33)*. Differential gene expression to determine the significantly differentially expressed genes between patients assigned to each cluster. A schematic of this data analysis pathway is shown in Figure 1.

Comparisons of relative immune mediator levels between subtypes were calculated using unpaired Student’s t test or Wilcoxon rank-sum test for non-parametric data. The associations between endotypes, clinical feature and outcomes was studied using logistic regression, the results of which were expressed as odd ratios with 95% confidence intervals. 30-day mortality was analyzed using Kaplan-Meier curves and cox-proportional hazards. Results were expressed as hazard ratios with 95% confidence intervals. P-values were corrected for multiple testing, if required, and considered significant if p < 0.05. Receiver operating curve statistics and confusion matrices were used to assess model fits on unseen data. All analysis was performed using R (version 3.6.0, R Core Team).

## Data Availability

Clinical data corresponding to raw and normalized microarray data are available in GEO, Data Accession Code GSE111368.

## Acknowledgments

Thank others for any contributions.

## Author contributions

CS, ERC, AIB and RCC conceived of the project and provided resources. PJMO and JD collected the data as part of the MOSAIC study. RS and AT analyzed the data. CS and RS drafted the manuscript. All authors reviewed and edited the manuscript prior to submission.

